# A predictive model for hospitalization and survival to COVID-19 in a retrospective population-based study

**DOI:** 10.1101/2022.03.02.22271552

**Authors:** Alejandro Cisterna-García, Antonio Guillén-Teruel, Marcos Caracena, Enrique Pérez, Fernando Jiménez, Francisco J. Francisco-Verdú, Gabriel Reina, Enrique González-Billalabeitia, José T. Palma, Álvaro Sánchez, Juan A. Botía

**Affiliations:** Departamento de Ingeniería de la Información y las Comunicaciones, Universidad de Murcia, Spain; Departamento de Informática, Servicio Murciano de Salud, Comunidad Autónoma de la Región de Murcia, Spain; Servicio de Microbiología. Clínica Universidad de Navarra, Pamplona, Spain; Department of Medical Oncology. Hospital Universitario 12 de Octubre, Madrid, Spain; Department of Biochemistry and Molecular Biology-A, Faculty of Biology, Regional Campus of International Excellence “Campus Mare Nostrum”, University of Murcia, Campus Espinardo, E-30100, Murcia, Spain; Murcia Biomedical Research Institute (IMIB-Arrixaca), 30120, Murcia, Spain; Universidad Católica San Antonio de Murcia-UCAM, Murcia, Spain; Department of Neurodegenerative Disease, Institute of Neurology, University College London, London, United Kingdom

## Abstract

The severe acute respiratory syndrome coronavirus (SARS-CoV-2) causing coronavirus disease 2019 (COVID-19) is highly transmissible and has been responsible for a pandemic associated with a high number of deaths. The clinical management of patients and the optimal use of resources are two important factors in reducing this mortality, especially in scenarios of high incidence. To this end, it is necessary to develop tools that allow early triage of patients with the minimal use of diagnostic tests and based on readily accessible data, such as electronic medical records. This work proposes the use of a machine learning model that allows the prediction of mortality and risk of hospitalization using simple demographic characteristics and comorbidities, using a COVID-19 dataset of 86867 patients. In addition, we developed a new method designed to deal with data imbalance problems. The model was able to predict with high accuracy (89-93%, ROC-AUC = 0.94) the patient’s final status (expired/discharged) and with medium accuracy the risk of hospitalization (71-73%, ROC-AUC = 0.75). These models were obtained by assembling and using easily obtainable clinical characteristics (2 demographic characteristics and 19 predictors of comorbidities). The most relevant features of these models were the following patient characteristics: age, sex, number of comorbidities, osteoarthritis, obesity, depression, and renal failure.

## Introduction

The virus responsible for Coronavirus disease 2019 (COVID-19), the severe acute respiratory syndrome coronavirus 2 (SARS-CoV-2), is a highly transmissible and pathogenic betacoronavirus that appeared in late 2019 in Wuhan, China ^1^. As of February 2022, it has had a tragic effect on the world’s population health resulting in more than 5,8 million deaths and 412 million cases worldwide, becoming the most important global health crisis since the era of the influenza pandemic of 1918 ^2,3^. The symptoms of COVID-19 are wide and may include fever, cough, fatigue, gastrointestinal issues, sore throat, anosmia, hyposmia, and neurological symptoms ^4–7^. Some of these symptoms can persist after recovery, particularly fatigue and dyspnea ^8^. The mortality rate of COVID-19 worldwide is around 1,5% since the pandemic began until February 2022 according to the World Health Organization (WHO). Although, we are aware of the existence of people more likely to develop a critical illness and eventually die ^9^. In addition, it has been proven that vaccines reduce COVID-19 death rates ^10,11^. For instance, the latest Centers from Disease Control and Prevention (CDC) release from the United States concluded that unvaccinated people have more COVID-19-associated death risk (mortality rate is around 1.39%) than fully vaccinated people (mortality rate is around 0.78%) with or without booster doses ^12^. Due to the high contagiousness and rapid spread of SARS-CoV-2, many countries have to manage intense periods of the disease, which are known as waves ^13^. In these periods, hospital resources, intensive care unit (ICU) capacity and healthcare system saturation can contribute to the increase in case fatality ^14^. Hence, clinical management of patients, quick risk stratification strategies, and optimization of the use of resources are important to reduce the fatality rate ^15,16^.

Electronic medical records (EHR) are one of the main resources to improve the way we approach patient management and move toward a more efficient triage of COVID-19 patients. Thus, patient demographic and health data available through healthcare systems have been used for COVID-19 patient prognosis and evolution through the use of semi-automated Artificial Intelligence (AI) systems. For example, a machine learning-based XGBoost model has been developed to predict patient mortality rates more than 10 days in advance with an accuracy of about 90%, using three biomarkers as main indicators for predicting COVID-19 prognosis, lactate dehydrogenase (LDH), high-sensitivity C-reactive protein (hs-CRP), and lymphocyte count ^17^. A higher ROC-AUC (0.96) was obtained with clinical data of patients at admission using four machine learning methods including logistic regression, support vector machine, decision tree with gradient boosting, and neural network ^18^. Using LASSO and a predictive equation with binary logistic regression based on pre-existing comorbidities and demographic data it was concluded that these variables demonstrated a good ability to discriminate severe from non-serious outcomes using only this historical information with an AUC of 0.76 ^19^. A further study developed models based on machine learning with different techniques, LASSO, novel univariate and pairwise, but concluded that no model was able to outperform a model based solely on age, where age had an AUC of 0.85 and balanced accuracy of 0.77 ^20^. Another model was able to predict the risk of hospital/ICU admission and death already at diagnosis with a ROC-AUC of 0.902 by focusing only on a limited number of comorbidities and demographic variables, such as age, sex, and BMI ^21^. In all of the above cases, the healthcare dataset is biased, as individuals with the most severe disease episodes are in the minority, so a supervised machine learning approach aimed at modeling them will suffer from imbalance. Moreover, the predictors considered for such studies are difficult to obtain. For example, LDH, albumin (ALB), blood urea nitrogen (BUN), hs-CRP, and lymphocytes require the use of blood or urea tests; or else the taking of measurements with specific physical devices, as is the case for BMI, temperature, and oxygen saturation. Therefore, these models are far from being realistically usable for early triage of patients in times of emergency oversaturation.

Our study presents a technique specially designed to address unbalanced problems applied to streamlining and improving the triage of COVID-19 patients according to their age, sex, and comorbidities, based on data readily available within the Regional Health System of the Region of Murcia, located in southeastern Spain. This allows for a local study in which data are organized in five different sources, including information on medical history, hospitalization services, symptoms, vital signs, treatments performed on more than 100,000 patients with COVID-19 with diagnosis dates ranging from January 4, 2020, to February 4, 2021, most of them prior vaccination (the COVID-19 vaccination program started in Spain on December 27, 2020) (See data collection). The technique to deal with unbalance consists of dividing the original problem into *p* subproblems, where each one will have a perfectly balanced data set associated with it, formed by samples from the original set. Using this reasoning, it is possible to build an ensemble logistic regression model that allows obtaining ROC-AUC of 0.94 to predict the final condition of the patient (discharge or expired) similar to complex models that combine several machine learning methods, and similar or higher than those that combine much more complex data based on laboratory or care techniques.

## Methods

### Study design and participants

Patients who were diagnosed with COVID-19 consecutively enrolled between January 4, 2020, and February 4, 2021, comprised our cohort. A confirmed case with COVID-19 is defined as a positive result of antigen test or real-time reverse-transcriptase polymerase-chain-reaction (RT-PCR) assay for nasal and pharyngeal swab specimens. Patients with incomplete records in the table “stratification of patients”, with active disease or lost to follow-up were excluded. The data in our analysis and models included 86867 confirmed COVID-19 patients (Figure 1). Features included in our study derived from the database “stratification of patients” which includes information for each patient about age, sex, diabetes mellitus, dementia, obesity, heart failure, chronic obstructive pulmonary disease (COPD), asthma, arterial hypertension, depression, ischemic cardiomyopathy, stroke, renal insufficiency, cirrhosis, osteoporosis, osteoarthritis, arthritis, Acquired Immune Deficiency Syndrome (VIH), and chronic pain.

**Figure 1.**
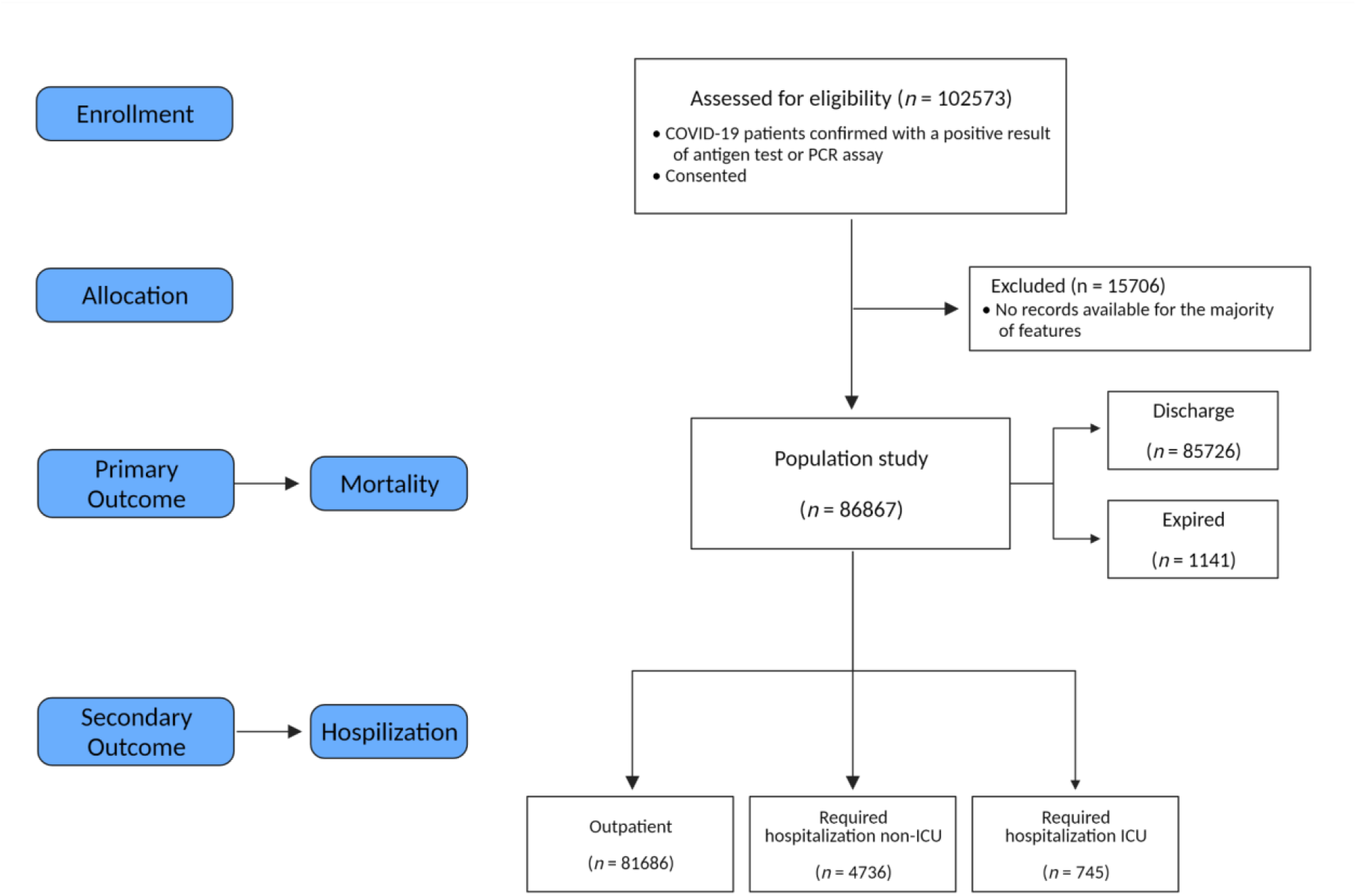
Consort diagram.

### Data collection

The COVID-19 epidemiological and clinical data were collected and derived from the EHR by the Regional Health System in Murcia (SMS), Spain. The patient database is arranged into five tables. The first table “stratification of patients” includes age, gender, hospital, and primary care team assigned to the patient, admission information, and final condition (i.e., the patient is cured or deceased), information about comorbidities, number of chronic pathologies, and number of affected systems, as well as the risk stratum, among other information, of a total of 102573 patients who were diagnosed with COVID-19. This is the main table on which the predictive models were developed. The second table “hospital dispensing drugs” contains information about the treatment of the patients who were hospitalized during their illness for COVID-19. This table includes the drugs administered, the dates on which they were taken, as well as the amount in which they were administered to 9165 patients who are present in the “stratification of patients” table. The third table “admissions and stays” contains information about inpatient stay services for 8356 patients in the “patient stratification” relation. Data about internal medicine, ICU, etc., and lengths of stay in each service for 8356 patients have passed during their hospitalization due to COVID-19 in the hospitals of the region. The fourth table “Selene medical record” contains information on vital signs, COVID-19 treatment, among other information, and the length of stay in each department of 7524 patients in the table “stratification of patients”. Some of the information collected in this relation is oxygen saturation, heart rate, body temperature, systolic and diastolic blood pressure, among others. The fifth table “OMI medical history” contains information about various symptoms, presented by 89769 patients in the database “stratification of patients”, such as headache, fever, dizziness, vomiting, among others. The relation “stratification of patients” is the central axis of the study, as it is the one that contains the information about the basic information necessary for the patients to know their clinical profile. The other databases contain additional useful information about the patients in this first database.

### Prediction models

We use machine learning to develop models about questions of interest: what the final condition of the subject will be and whether the individual will be hospitalized. We want to be able to answer these questions when the patient has just been diagnosed, therefore the only information that we can use for that is obtained in a medical review or the previous information available in the EHR of the patient. In this study, what we have at our disposition is a table that contains information on the age, sex, comorbidities, hospitalization status, and final outcome of the patients. The available datasets for both questions are highly imbalanced, as there are far fewer expired individuals (1141) than discharged patients (85726), and there are more outpatients (81386) than hospitalized (non-ICU 4736 and ICU 745). To address these issues, we propose a new imbalance-aware and ensemble-based machine learning method. It is called IPIP (Identical Partitions for Imbalance Problems). First, we hold out 20% of the data from the minority class and the same number of samples from the majority class to create a test set. The rest goes into a train set. We then divide the training data set into *p* perfectly balanced data sets. For each, we create a basic ML model. All models go into an ensemble whose response aggregation is a simple majority. IPIP selects *p* depending on *n* (number of samples). Higher values of *n* lead to lower values of *p*. For this particular dataset, 75% of the minority class samples of the train data leads to p= 7 perfectly balanced subsets. Within each *p* iteration, it randomly splits into train and test datasets and generates a model with training data and testing with test data. If the new model improves the overall quality of the ensemble, it is added. If not, it randomly samples and tries again up to a max number of attempts. We use the test set to evaluate the candidate ensemble for improvement. When doing inference, the classifier predicts an observation as a member of the majority class when at least 75% of the models classified as negative (majority class) will be classified as negative. The final ensemble’s inference in production mode is generated by evaluating each one of the models that compose the final ensemble, if 50% of them classify a sample as negative (majority class), the final model classifies it as negative.

We created two IPIP models to address each modeling question. One with a baseline algorithm, logistic regression, and the other with random forests ^22^ as basic models based on the Caret R package ^23^. All models were evaluated using five-fold cross-validation. Different values of the number of decision trees for the random forest models were tested, and we decided that each random forest model used 200 decision trees, where the impurity is the variable importance mode, which is the Gini index for classification. A tune grid was created to choose the better minimal node size of the trees (1, 11, or 21) and the number of variables to possibly split into each node (1, 4, 7, 10, 13, 16, or 19). To decide whether a basic model improves the ensemble or not, we relied on Cohen’s Kappa metric ^24^. That is, if adding a basic model to the set of basic models trained for a specific perfectly balanced subset improved the Kappa in the evaluation on the available test set of the new set of basic models concerning the previous Kappa values on the same evaluation set, we added that basic model to that set of basic models, otherwise it was discarded. We also obtained the following metrics: balanced accuracy, negative predictive value (NPV), positive predictive value (PPV), sensitivity, and specificity. In addition, we computed the Receiver Operating Characteristics Area Under the Curve (ROC-AUC) for the final ensemble model.

### Statistical Analysis

Continuous data are reported as median with interquartile range (IQR), and categorical data are expressed as percentages (%). We also used odds ratio (OR) and 95% CIs. We adjusted OR by gender and age. Differences between groups were tested using the Mann-Whitney U for numerical variables, and χ2 test or Fisher’s exact test was used to test significance for categorical data. Statistical analysis was performed by using R (version 3.6.3) with P-values significance threshold of 0.05.

## Results

### Description and differences of the different types of COVID-19 patients in our dataset

The exploratory analysis of the data from 86867 COVID-19 patients in a region located in the southeast of Spain (Region of Murcia) allowed stratifying the database obtained by age, sex, and specific comorbidities (Table 1), following the flow chart for the cohort shown in Figure 1. Among the cases studied, 93.7% were outpatients (N=81386), 5.4% were hospitalized non-ICU (N=4736) and less than 0.85% were patients admitted to the ICU (N=745). The most common symptoms among patients were cough (49.9%), followed by headache (38.3%) and myalgia (36%) (Supplementary Table S1). Using the data in Table 1 we can identify the different prototypes of patients with COVID-19.

**Table 1.** Demographic characteristics, comorbidities, and final outcome of different types of COVID-19 patients.

| Characteristics | Outpatient | Hospitalized (non-ICU) | ICU |
| --- | --- | --- | --- |
| Number of individuals (N) | 81386 | 4736 | 745 |
| Age median<br>(IQR) | 38.00<br>(22.00, 52.00) | 62.00<br>(47.00, 79.00) | 62<br>(52.00, 71.00) |
| **Gender** |  |  |  |
| Male<br>(%) | 38200<br>(46.94%) | 2450<br>(51.73%) | 524<br>(70.34%) |
| Female<br>(%) | 43186<br>(53.06%) | 2286<br>(48.27%) | 221<br>(29.66%) |
| **Comorbidities** |  |  |  |
| Number of Chronic diseases median<br>(IQR) | 2.00<br>(1.00, 4.00) | 5.00<br>(2.00, 8.25) | 5<br>(2.00, 8.00) |
| Number of systems affected median<br>(IQR) | 2.00<br>(1.00, 3.00) | 4.00<br>(2.00, 5.00) | 3<br>(2.00, 5.00) |
| Asthma<br>(%) | 7,032<br>(8.64%) | 398<br>(8.40%) | 70<br>(9.40%) |
| Obesity<br>(%) | 7,516<br>(9.24%) | 1,008<br>(21.28%) | 220<br>(29.53%) |
| Diabetes mellitus<br>(%) | 5,212<br>(6.40%) | 1,197<br>(25.27%) | 214<br>(28.72%) |
| Heart failure<br>(%) | 646<br>(0.79%) | 343<br>(7.24%) | 33<br>(4.43%) |
| COPD<br>(%) | 1,059<br>(1.30%) | 350<br>(7.39%) | 46<br>(6.17%) |
| Arterial hypertension<br>(%) | 12,210<br>(15.00%) | 2,216<br>(46.79%) | 356<br>(47.79%) |
| Depression<br>(%) | 7,345<br>(9.02%) | 800<br>(16.89%) | 103<br>(13.83%) |
| HIV<br>(%) | 121<br>(0.15%) | 10<br>(0.21%) | 3<br>(0.40%) |
| Ischemic cardiomyopathy<br>(%) | 1,349<br>(1.66%) | 420<br>(8.87%) | 75<br>(10.07%) |
| Stroke<br>(%) | 961<br>(1.18%) | 353<br>(7.45%) | 37<br>(4.97%) |
| Renal insufficiency<br>(%) | 1,267<br>(1.56%) | 515<br>(10.87%) | 66<br>(8.86%) |
| Cirrhosis<br>(%) | 1,672<br>(2.05%) | 256<br>(5.41%) | 57<br>(7.65%) |
| Osteoporosis<br>(%) | 2,453<br>(3.01%) | 431<br>(9.10%) | 51<br>(6.85%) |
| Osteoarthritis<br>(%) | 5,212<br>(6.40%) | 854<br>(18.03%) | 121<br>(16.24%) |
| Arthritis<br>(%) | 1,111<br>(1.37%) | 132<br>(2.79%) | 21<br>(2.82%) |
| Dementia<br>(%) | 859<br>(1.06%) | 296<br>(6.25%) | 9<br>(1.21%) |
| Chronic pain<br>(%) | 39<br>(0.05%) | 9<br>(0.19%) | 4<br>(0.54%) |
| **Outcome** |  |  |  |
| Discharge<br>(%) | 81,132<br>(99.69%) | 4,082<br>(86.19%) | 512<br>(68.72%) |
| Expired<br>(%) | 254<br>(0.31%) | 654<br>(13.81%) | 233<br>(31.28%) |

Continuous data are reported as median with interquartile range (Q3-Q1), and categorical data are expressed as percentages (%). We also used odds ratio (OR) and 95% CIs. COPD is a chronic obstructive pulmonary disease and HIV is Acquired Immune Deficiency Syndrome. Expired (report only when the patient dies).

Three types of patients were identified from the exploratory analysis (Table 1), the COVID-19 outpatient prototype was a 38-year-old female (IQR: 22-52) (53.06% of our outpatients are female), with 2 chronic pathologies and 2 affected systems, whose most common pathologies or comorbidities were arterial hypertension (15.00%), followed by obesity (9.24%), depression (9.02%), and asthma (8.64%). In contrast, the prototypical COVID-19 hospitalized non-ICU patient was a 62-year-old male (IQR: 47-79) (51.73% were men), with 5 chronic pathologies, with 4 affected systems, and with more frequent pathologies or comorbidities such as arterial hypertension (46.79%), diabetes mellitus (25.27%), obesity (21.28%), osteoarthritis (18.03%) and depression (16.89%). Finally, the profile of the patient admitted to the ICU was a 62-year-old male (IQR: 52-71) (70.34% of ICU patients were male), with 5 chronic pathologies, 3 affected systems, and whose most frequent pathologies or comorbidities were arterial hypertension (47.79%), followed by obesity 29.53%, diabetes mellitus 28.72%, and osteoarthritis (18.03%). COVID-19 Patients in the ICU had almost twice the possibility of dying than those hospitalized not admitted to the ICU (31.28% vs. 13.81%), far removed from that presented by outpatients (0.31%).

To further study the differences between discharged COVID-19 patients (survivors) and those who deceased (non-survivors), the data in Table 1 were reorganized in Table 2. The prototype of the surviving patient was a 39-year-old female (IQR: 23-53) (52.72% are female), with 2 chronic pathologies, 2 affected systems, and with more frequent pathologies or comorbidities similar to those previously described for the non-hospitalized patients. In contrast, the profile of the non-surviving patient was clearly different and represented by an 83-year-old man (IQR: 75-88) (56.00% were men), with 8 chronic pathologies, 5 affected systems, and whose prevalent pathology or comorbidity was arterial hypertension (75.64%), far from the subsequent ones such as diabetes mellitus (42.33%), obesity (29.36%), osteoarthritis (27.93%) and depression (23.31%). In addition, three variables were highly relevant to the patient’s final status (Figure 2). Thus, the older the patient *t*(1237) = 116.9, *p* < 2.2×10-16, the greater the number of chronic pathologies *t*(1151) = 42.15, *p* < 2.2×10-16 and the greater the number of affected systems *t*(1163) = 47.2, *p* < 2.2×10-16, the greater the probability of death (Figure 2). A similar distribution of the above variables was observed when the population was divided into the three initial groups (outpatients, hospitalized non-ICU, and ICU) (Supplementary Figure S1). The relationship between gender and patient status was significant (X^2^ _(1, N = 86867)_ = 34.33, *p* = 4.64×10-9). Thus, men were more likely to die than women (Table 2, Supplementary Figure 2), and that the highest risk comorbidities or pathologies were renal insufficiency (OR=1.90, 95% CI: 1.61, 2.24), heart failure (OR=1.85; 95% CI: 1.53, 2.23), stroke (OR=1.84 95% CI: 1.54, 2.20), dementia (OR=1.78; 95% CI: 1.50, 1.81) and ischemic cardiomyopathy (OR=1.58; 95% CI: 1.33, 1.88). However, there was no significant relationship between asthma, osteoarthritis, and osteoporosis with COVID-19– related death.

**Figure 2.**
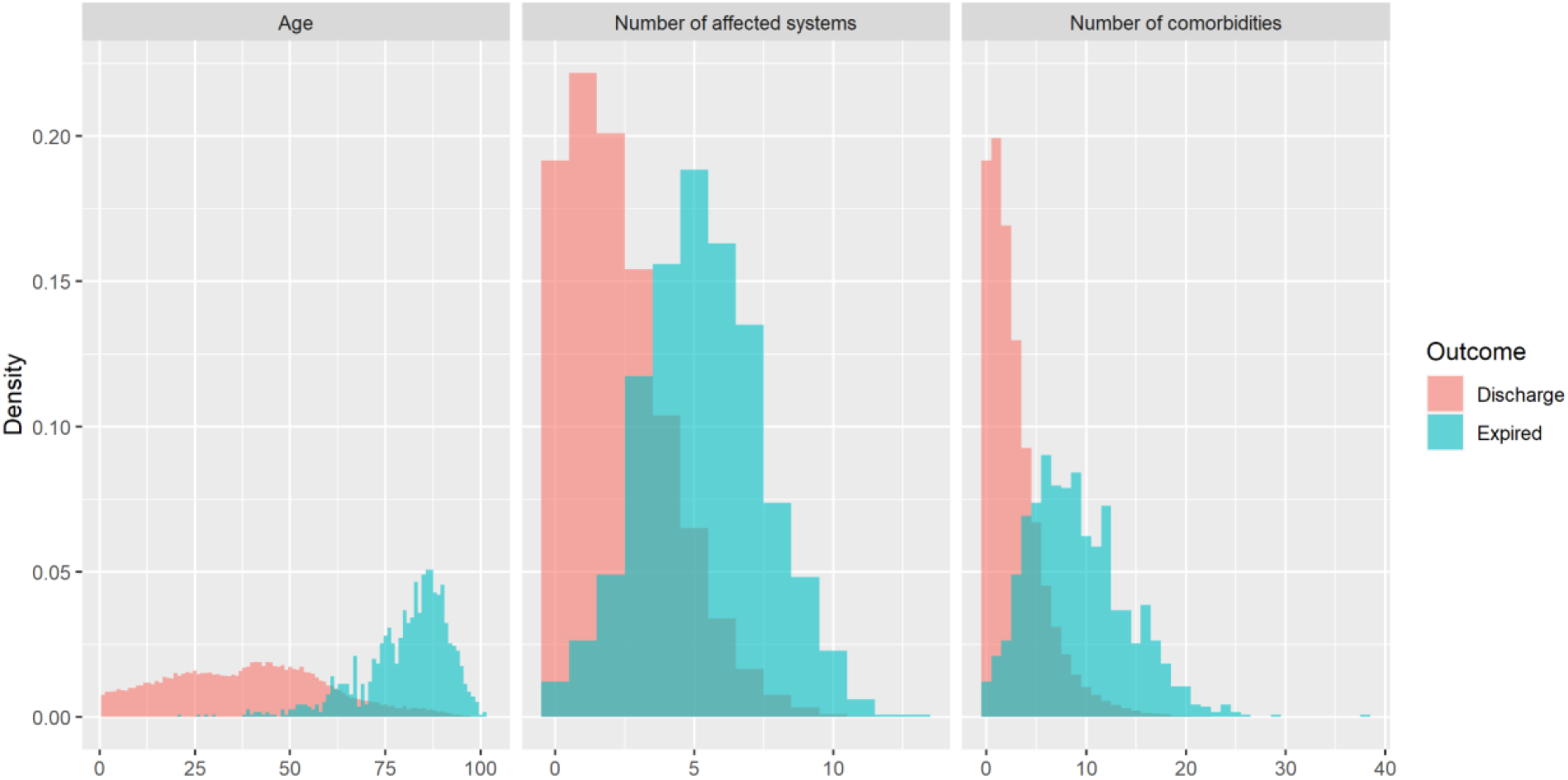
Distribution of age, the number of affected systems, and the number of comorbidities regarding the final patient outcome. We clearly see different distributions of these variables between discharge and expired patients.

**Table 2.** Demographic characteristics and comorbidities of discharge and expired COVID-19 patients.

| Characteristics | Discharge<br>(survival) | Expired<br>(non- survival) | OR<br>(95% CIs) |
| --- | --- | --- | --- |
| Number of individuals (N) | 85726 | 1141 | - |
| Age median<br>(IQR) | 39.00<br>(23.00 53.00) | 83<br>(75.00 88.00) | - |
| **Gender** |  |  |  |
| Male<br>(%) | 40535<br>(47.28%) | 639<br>(56.00%) | 2.41<br>(2.11 2.75) |
| Female<br>(%) | 45191<br>(52.72%) | 502<br>(44.00%) | - |
| **Comorbidities** |  |  |  |
| Number of Chronic diseases median<br>(IQR) | 2.00<br>(1.00 4.00) | 8<br>(5.00 12.00) | - |
| Number of systems affected median<br>(IQR) | 2.00<br>(1.00 3.00) | 5<br>(4.00 7.00) | - |
| Asthma<br>(%) | 7403<br>(8.64%) | 97<br>(8.50%) | 1.13<br>(0.90 1.42) |
| Obesity<br>(%) | 8409<br>(9.81%) | 335<br>(29.36%) | 1.57<br>(1.36 1.81) |
| Diabetes mellitus<br>(%) | 6140<br>(7.16%) | 483<br>(42.33%) | 1.55<br>(1.36 1.77) |
| Heart failure<br>(%) | 823<br>(0.96%) | 199<br>(17.44%) | 1.85<br>(1.53 2.23) |
| COPD<br>(%) | 1290<br>(1.50%) | 165<br>(14.46%) | 1.49<br>(1.23 1.81) |
| Arterial hypertension<br>(%) | 13919<br>(16.24%) | 863<br>(75.64%) | 1.31<br>(1.12 1.53) |
| Depression<br>(%) | 7982<br>(9.31%) | 266<br>(23.31%) | 1.24<br>(1.06 1.44) |
| HIV<br>(%) | 134<br>(0.16%) | 0<br>(0.00%) | - |
| Ischemic cardiomyopathy<br>(%) | 1631<br>(1.90%) | 213<br>(18.67%) | 1.58<br>(1.33 1.88) |
| Stroke<br>(%) | 1145<br>(1.34%) | 206<br>(18.05%) | 1.84<br>(1.54 2.20) |
| Renal insufficiency<br>(%) | 1591<br>(1.86%) | 257<br>(22.52%) | 1.90<br>(1.61 2.24) |
| Cirrhosis<br>(%) | 1923<br>(2.24%) | 62<br>(5.43%) | 1.41<br>(1.07 1.86) |
| Osteoporosis<br>(%) | 2758<br>(3.22%) | 177<br>(15.51%) | 1.06<br>(0.88 1.29) |
| Osteoarthritis<br>(%) | 5868<br>(6.85%) | 319<br>(27.96%) | 1.03<br>(0.89 1.20) |
| Arthritis<br>(%) | 1214<br>(1.42%) | 50<br>(4.38%) | 1.44<br>(1.05 1.98) |
| Dementia<br>(%) | 911<br>(1.06%) | 253<br>(22.17%) | 1.78<br>(1.50 2.12) |
| Chronic pain<br>(%) | 50<br>(0.06%) | 2<br>(0.18%) | - |

Continuous data are reported as median with interquartile range (Q3-Q1) and categorical data are expressed as percentages (%). We also used odds ratio (OR) and 95% confidence intervals CIs. COPD is a chronic obstructive pulmonary disease and HIV is Acquired Immune Deficiency Syndrome. Expired (report only when the patient dies).

### ML models

Several machine learning models were developed: 1) to predict the patient’s final condition and 2) to predict which patient will need to be hospitalized. The training dataset (85476 surviving and 891 non-surviving patients) was used to train the model to predict the final patient’s condition and the test dataset (500 patients; 250 patients from each class) was used to evaluate this model. To realistically evaluate the model, 101 test sets were created with the same proportion as the initial set (1/75), i.e., for each deceased COVID-19 patient, 75 surviving patients were taken.

Two machine learning algorithms (Random Forest and Logistic Regression) were evaluated with or without IPIP, a method to deal with unbalanced data (see Methods). The accuracy and Cohen’s Kappa obtained on the test data set for the ensemble models (Figure 3) showed that the IPIP model with logistic regression (LR-IPIP) obtained the best results regarding the final condition of the patient. This LR-IPIP model combined the result of the ensemble of 17 logistic regression models. The patient’s final condition can be predicted with the LR-IPIP model with a balanced accuracy between 0.89 and 0.93 (Table 3) for the imbalanced datasets versus 0.91 for the balanced datasets, balanced datasets are the ones usually used in the literature and which give a higher Cohen’s Kappa coefficient (0.82 vs. 0.18). In addition, the ROC-AUC of this model for the unbalanced datasets was 0.937 (Supplementary Figure S3A). The most important factors determining the patient’s final condition (Importance Features) obtained by this RL-IPIP model were firstly age (FI: 1.0), followed by sex (FI: 0.34), osteoarthritis (FI: 0.21), obesity (FI: 0.15) and the number of affected systems (FI: 0.14) (Figure 4A, Supplementary Figure S4).

**Figure 3.**
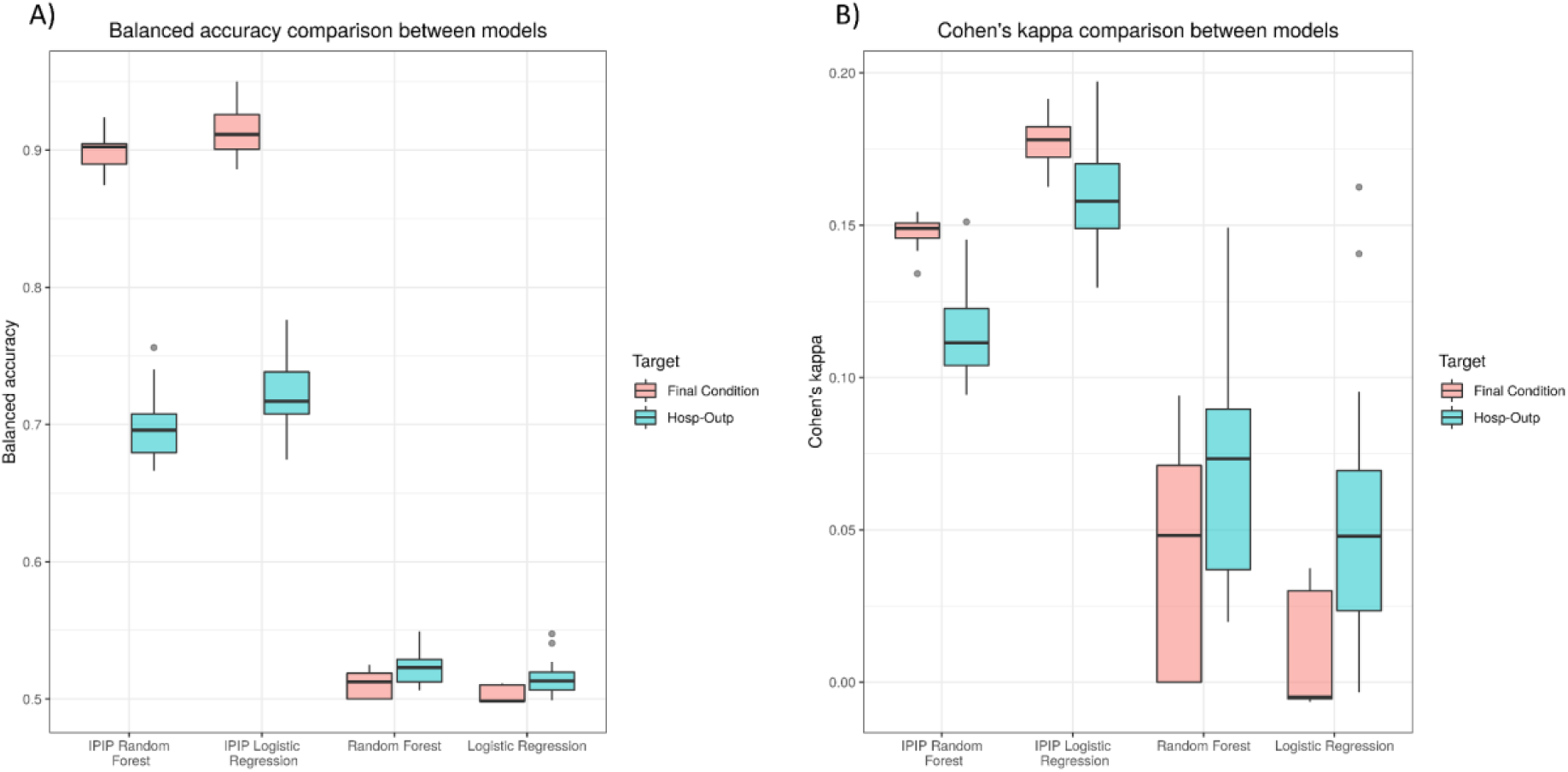
A) Accuracy boxplots obtained in the test of the models trained for each problem. B) Cohen’s Kappa coefficients boxplots were obtained in the test of the models trained for each problem.

**Table 3.** Metrics obtained in the test dataset using the best training model.

| Metrics | Expired/Discharge |  | Hospitalized/Outpatient |  |
| --- | --- | --- | --- | --- |
|  | Unbalanced tests | Balanced test | Unbalanced tests | Balanced test |
| **Balanced accuracy** | 0.91 (0.89, 0.93) | 0.91 | 0.72 (0.71, 0.73) | 0.72 |
| **Cohen's Kappa** | 0.18 (0.17, 0.19) | 0.82 | 0.16 (0.15, 0.17) | 0.44 |
| **Sensitivity** | 0.93 (0.88, 0.97) | 0.92 | 0.72 (0.70, 0.74) | 0.71 |
| **Specificity** | 0.90 (0.89, 0.90) | 0.90 | 0.73 (0.72, 0.73) | 0.73 |
| **Positive predictive value (PPV)** | 0.11 (0.10, 0.12) | 0.90 | 0.15 (0.14, 0.15) | 0.72 |
| **Negative predictive value (NPV)** | 1 (0.99, 1) | 0.92 | 0.98 (0.97, 0.98) | 0.72 |

*Balanced tests are sets with the same number of patients of each class. Unbalanced tests are sets with the same proportion of classes as the original dataset, where the number of examples for each class label is not balanced. Those sets are made up by a chosen number of samples of patients for each class. For unbalanced tests we report the mean (95% CIs) of each metric, and for balanced tests we report the value of each metric.*

**Figure 4.**
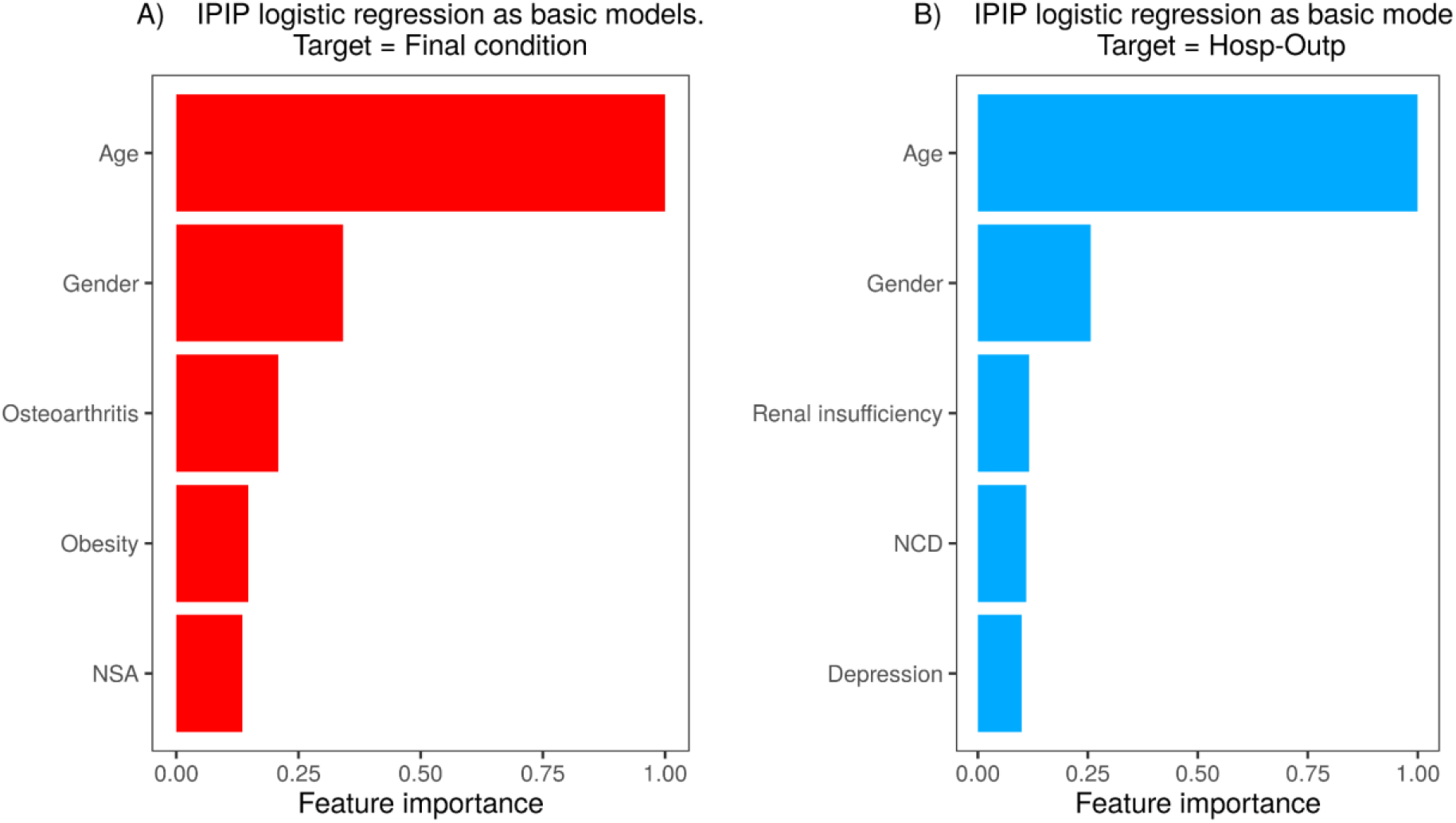
Feature importance for the final models. NSA is the number of systems affected and NCD is the number of chronic diseases.

On the other hand, the model to predict which patient will need to be hospitalized was developed using a training dataset (4385 inpatients and 80290 outpatients), and a test dataset (2192 patients; 1096 patients from each class) was used to evaluate this model. In turn, this data was distributed in 25 test sets with the same ratio of inpatients/outpatients as in the initial set (1/15). Again, the model with the best results was the LR-IPIP made up of 13 logistic regression models (Figure 3). The need for hospitalization could be predicted with the RL-IPIP model with a balanced accuracy between 0.71 and 0.73 (Table 3) for the imbalanced datasets versus 0.72 for the balanced datasets. Similar to the other model, the Cohen’s Kappa coefficient is higher for the balanced dataset (0.44 vs. 0.16). In addition, the ROC-AUC of this model for the unbalanced datasets was 0.746 (Supplementary Figure S3B). Finally, the significance of the characteristics obtained in that model showed that age (FI: 1.0) was also the most relevant characteristic, followed by sex (FI: 0.26), renal insufficiency (FI: 0.12), number of chronic diseases (FI: 0.11) and depression (FI: 0.1) (Figure 4B, Supplementary Figure S4).

## Discussion

In this study we have analyzed the different COVID-19 patient types in Southeastern Spain (n=86867). In contrast to most COVID-19 studies that developed predictive models in the literature that handle less than 5000 patients ^17–21^. In addition, we have presented a technique specially designed to treat imbalance problems (IPIP), with which we have developed machine learning models to predict the final state of the patient and the need for hospitalization of those. We trained and evaluated the models with and without IPIP, which efficiently manages the imbalance in the data according to our results (Figure 3).

Regarding characterizing the different kinds of prototypical COVID-19 patients, in this region, the COVID-19 outpatient prototype is a 38-year-old woman, with 2 chronic pathologies whereas the hospitalized COVID-19 patient prototype is a 62-year-old man, with 5 chronic pathologies. We identified age, gender, and the number of comorbidities as important to distinguish between outpatient and hospitalized. Several studies have also found that hospitalized COVID-19 patients are more commonly older, male, and associated with more comorbidities such as obesity, diabetes mellitus, and hypertension ^25,26^. In addition, we could find statistically significant differences for age (*p* < 8.0×10-3), the number of comorbidities (*p* < 2.5×10-3), and gender (*p* < 2.2×10-16) between ICU and hospitalized non-ICU patients, although those differences are smaller than between outpatient and hospitalized. ICU patients were around a year younger than hospitalized non-ICU patients and had fewer comorbidities (Supplementary Figure S1). Therefore, we hypothesized that clinicians included patients more likely to survive in the ICU because of the limited number of available ICU slots or the risk of the male gender. We also detect even more differences for those features between survivors (discharge patients) and non-survivors (expired patients) (Figure 2). In our region, the discharge patient prototype is a 39-year-old woman, with 2 chronic pathologies while the expired patient prototype is an 83-year-old man, with 8 chronic pathologies. According to several studies, our results show that older patients are more likely to die ^27–29^, and also male patients are more likely to die (OR=2.41, 95% CI: 2.11, 2.75) (Table 2, Supplementary Figure 2) ^30,31^. When it comes down to comorbidities, we found that asthma, osteoporosis, and osteoarthritis are not associated with COVID-19–related death. A large number of studies report that patients with asthma are not at risk of severe COVID-19 ^32,33^. For osteoarthritis association with COVID-19–related death we found a study that reported similar OR=0.84 (95% CI: 0.65–1.08) ^34^. For osteoporosis, it is known that women are more at risk of developing osteoporosis than men ^35^. It seems that some particular kinds of osteoporosis complications are associated with more risk of COVID-19 exitus, however, this study did not adjust the risk by age and gender ^36^. The rest of the comorbidities evaluated in our study were associated with an increase in mortality risk. These comorbidities or pathologies are diabetes mellitus, dementia, obesity, heart failure, COPD, arterial hypertension, ischemic cardiomyopathy, stroke, renal insufficiency, cirrhosis, and arthritis. Several studies obtain the same results for those comorbidities ^34,37,38^. Regarding depression, in line with our results, a meta-analysis identified that depression is associated with more COVID-19–related death ^39^. All the results mentioned above are important to ensure that the characteristics and comorbidities of our population were not unique. In addition, we believe that due to the similarity with other COVID-19 studies our data could be useful to develop predictive models.

Since the beginning of the pandemic, there have been many studies that have reported some important clinical characteristics (predictors) for mortality in patients with COVID-19 through the development of ML-based models. Selected characteristics used as inputs for the development of these models included baseline data, clinical symptoms, associated comorbidity, and clinical indicators. However, these studies have two fundamental problems: the low number of patients due to the number of parameters studied greatly restricts the cohort and the strongly unbalanced data. To bridge these drawbacks, in this work we tested different ML models considering basic data easily accessible in an emergency care setting and based on clinical data from EHR to help during early patient triage. We definitely obtained promising results when predicting the patient’s final condition using the LR-IPIP model (0.91 balanced accuracy, ROC-AUC = 0.94). In terms of variable importance, ML detects Age (FI: 1.0), gender (FI: 0.34), osteoarthritis (FI: 0.21), obesity (FI: 0.15), and the number of systems affected (FI: 0.14) as the most important variables to predict exitus. The model also detected comorbidities such as renal insufficiency, dementia, diabetes mellitus, and COPD. These features are associated with more risk of COVID-19–related death according to our model. In a similar direction, these comorbidities are associated with severe clinical manifestations observed in older adult patients ^40,41^. Comorbidities such as cardiovascular disease, hypertension, and diabetes although are highly prevalent in older adults have been associated with worse outcomes in COVID-19 ^34,37,38^. Studies that rely on comorbidities to predict death based on ML usually rank age as one of the most influential variables ^42,43^, in fact, a meta-analysis with 611,583 patients demonstrates an age-related increase in mortality. Thus, the highest mortality occurs in patients >80 years, in whom it was 6 times higher than in younger patients ^44^. Similarly, gender is an important feature for several ML-based studies ^42,45^, our model identified that male patients are more likely to die, perhaps due to the distribution of our data (OR=2.41, 95% CI: 2.11, 2.75), which is in agreement with previous work ^30,31^. Similar to our model, another ML-based study identified obesity as an important feature ^46^. However, to the best of our knowledge, this is the first time that a model reports osteoarthritis as an important feature. The beta values in the ensemble model showed that osteoarthritis is associated with less risk of COVID-19–related death (Supplementary table S2). This might be in agreement with a study using UK biobank data (OR =0.84, 95% CI 0.65-1.08), although it is not statistically significant ^34^. In addition, the osteoarthritis distribution in our population is not statistically associated with the patient’s final condition. Note that, although we have no conclusive evidence on this, patients with osteoarthritis may be subjected to medication. Interestingly, we might think that medication could play a role in patients with osteoarthritis and COVID-19, however, Wong et al. reported that non-steroidal anti-inflammatory drugs (NSAIDs) medication is not associated with a higher risk of COVID-19 death for osteoarthritis patients ^47^. Dementia, together with the number of affected systems and the number of comorbidities, also appear among the most relevant characteristics, which is in agreement with the aforementioned factors in other studies, and in the case of dementia, with the results obtained from a cohort of 12863 individuals from the UK Biobank who lived in the community and were over 65 years of age (1814 individuals ≥ 80 years of age) were tested for COVID-19, where it was seen that all causes of dementia increased the risk of death related to COVID-19 ^48^. Regarding accuracy, our LR-IPIP model obtained a balanced accuracy between 89-93% (ROC-AUC = 0.94) in predicting the patient’s final condition. Accuracy was similar to or higher than others if we compare our results with several studies. For instance, Gao et al. reported an accuracy between 80.6-96.8% ^18^ which is a large confidence interval besides they used more complex clinical data points on admission. Chatterjee et al. reported a balanced accuracy of 72% ^20^, perhaps due to the low number of COVID-19 patients. Finally, another ML-based study was able to predict the risk of death already at diagnosis with a ROC-AUC of 0.902 ^21^.

The ability of the LR-IPIP model to decide the hospitalization of new patients was not as efficient (balanced precision = 0.72; ROC-AUC = 0.75). Regarding the importance of the variables, ML again found that age, gender, and the number of comorbidities were important. Among these, obesity reappears, and renal insufficiency and depression appear in a prominent place. Thus, it has been shown that acute renal failure is frequent among patients hospitalized for COVID-19 and that only 30% survived with the recovery of renal function at discharge ^49^.

In conclusion, this paper shows the analysis and development of predictive ML-based models with one of the largest COVID-19 datasets (n=86867) obtained from the health service of the Region of Murcia (Spain). In addition, the problem of class imbalance has been addressed by developing a new algorithm, called IPIP, which automatically deals with this problem. The model obtained allows predicting with high accuracy the final state of the patient, and with reasonable precision which patient will need to be hospitalized, simply by using the demographic data and comorbidities accessible at COVID-19 diagnosis by the clinicians. In fact, this LR-IPIP predictive model can be used, among other considerations, to prioritize triage of COVID-19 patients when health system resources are limited, as is often the case during different waves of COVID-19. To facilitate this prioritization of resources, both the corresponding web application and the predictive models are easily accessible in open repositories (GitHub), which will facilitate their adaptation to new datasets of future epidemic waves of this disease or other respiratory viruses in general.

## Data Availability

All data produced in the present study are available upon reasonable request to the authors and Murcia health service

## CODE AVAILABILITY

The functions and models required to reproduce the results are available at: https://github.com/antoniogt/ipip

## DATA AVAILABILITY

We developed a shiny app to share our model and to predict patient hospitalization https://alejandrocisterna.shinyapps.io/PROVIA

## Supplementary

**Supplementary Table S1.**
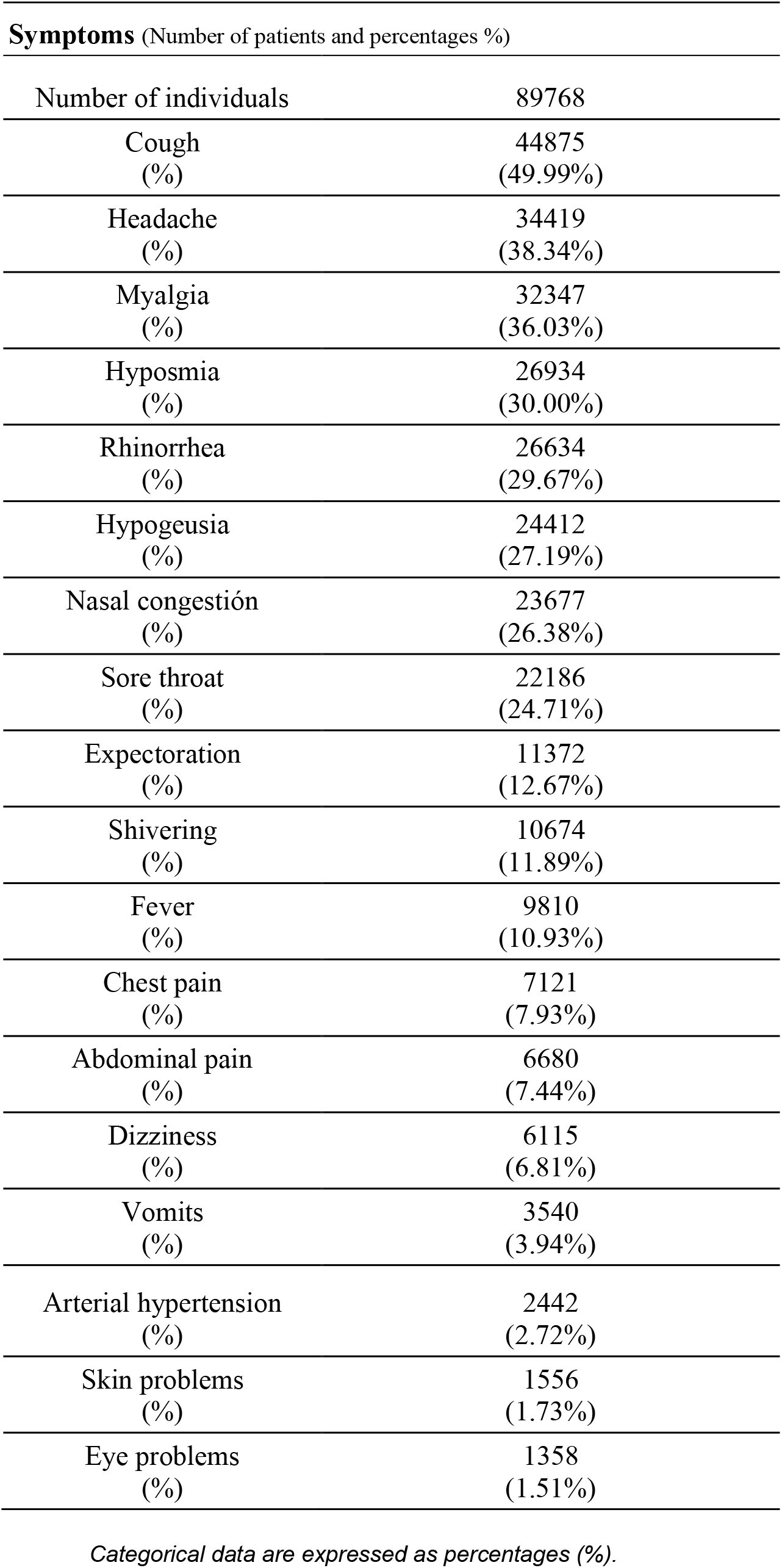
Symptoms and its frequency in COVID-19 patients.

**Supplementary Table S2.** Beta values (osteoarthritis) for each of 17 logistic regression models (Final condition ensemble).

| Features | Logistic Regression Model. Target: Final Condition |  |  |  |  |  |  |  |  |  |  |  |  |  |  |  |  |
| --- | --- | --- | --- | --- | --- | --- | --- | --- | --- | --- | --- | --- | --- | --- | --- | --- | --- |
|  | 1 | 2 | 3 | 4 | 5 | 6 | 7 | 8 | 9 | 10 | 11 | 12 | 13 | 14 | 15 | 16 | 17 |
| Osteoarthritis<br>– Yes | -0.164 | -1.109 | -0.529 | -0.897 | -1.782 | -1.112 | -1.092 | -0.334 | -0.822 | -0.912 | -0.573 | -0.584 | -1.279 | -0.978 | -0.558 | -0.729 | -1.193 |

**Supplementary Figure S1.**
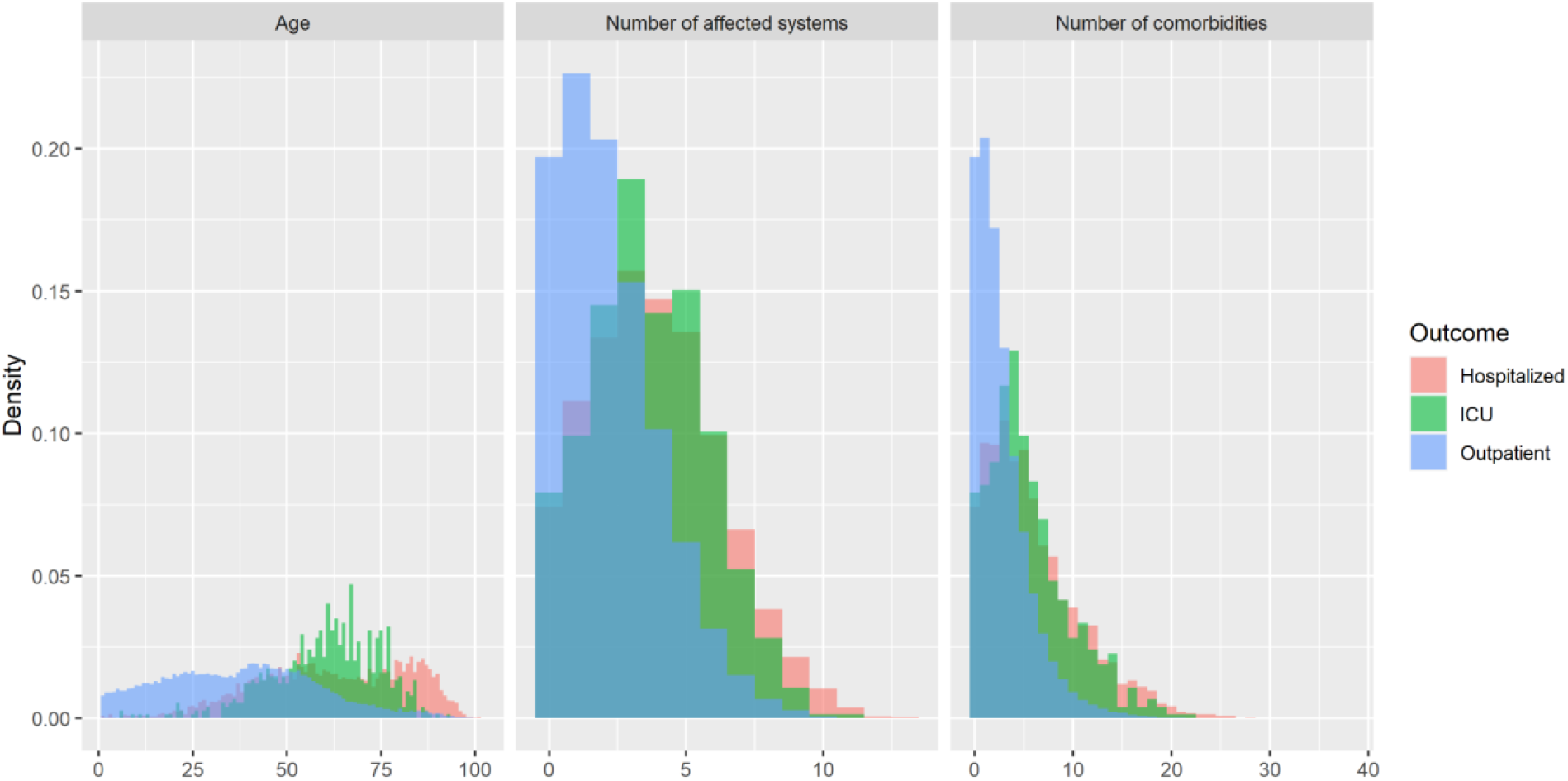
Distribution of age, the number of affected systems, and the number of comorbidities regarding the type of patient. We clearly see different distributions of these variables between outpatient and hospitalized (ICU and non-ICU) patients.

**Supplementary Figure S2.**
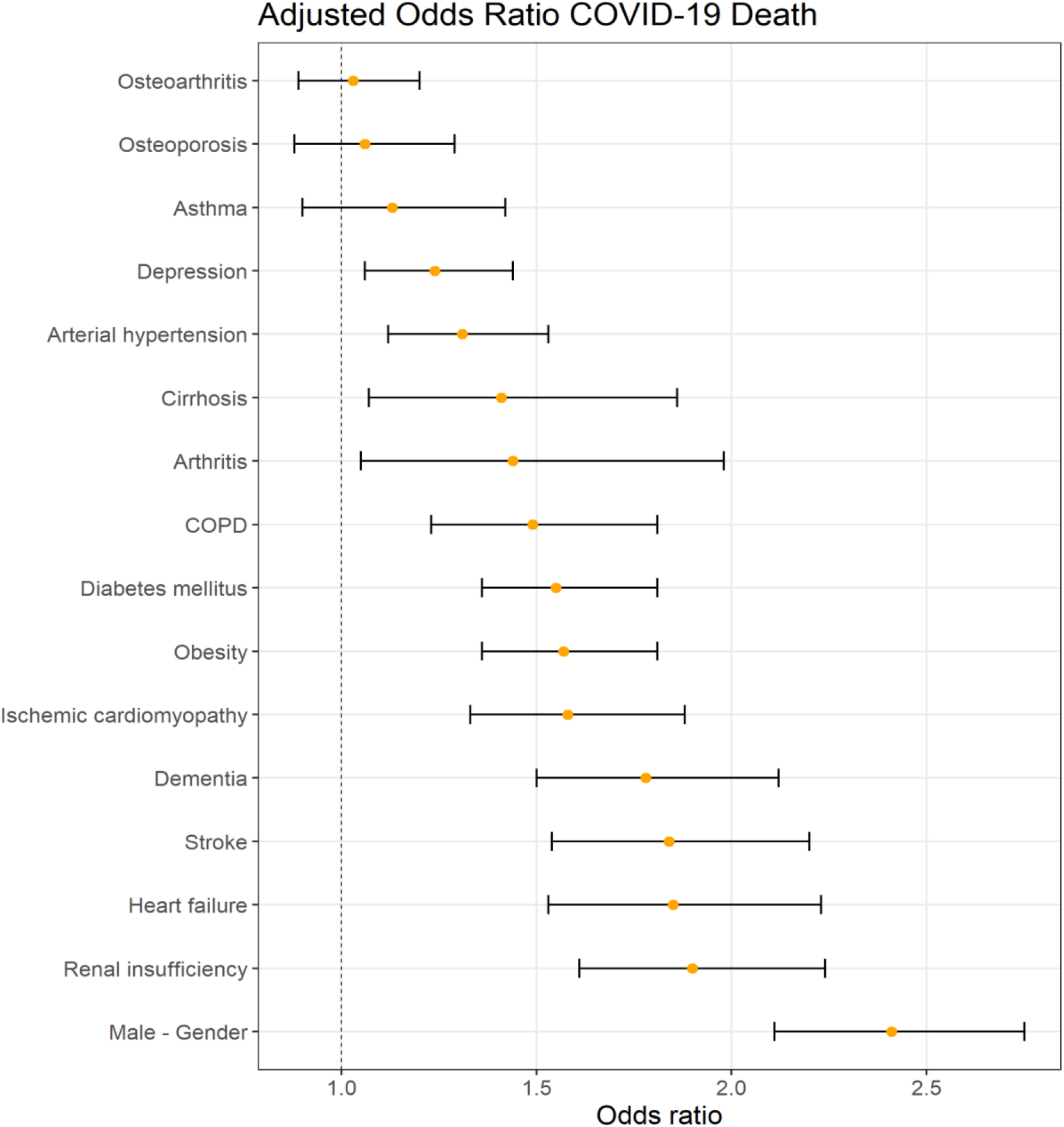
Adjusted Odds Ratio COVID-19 Death

**Supplementary Figure S3.**
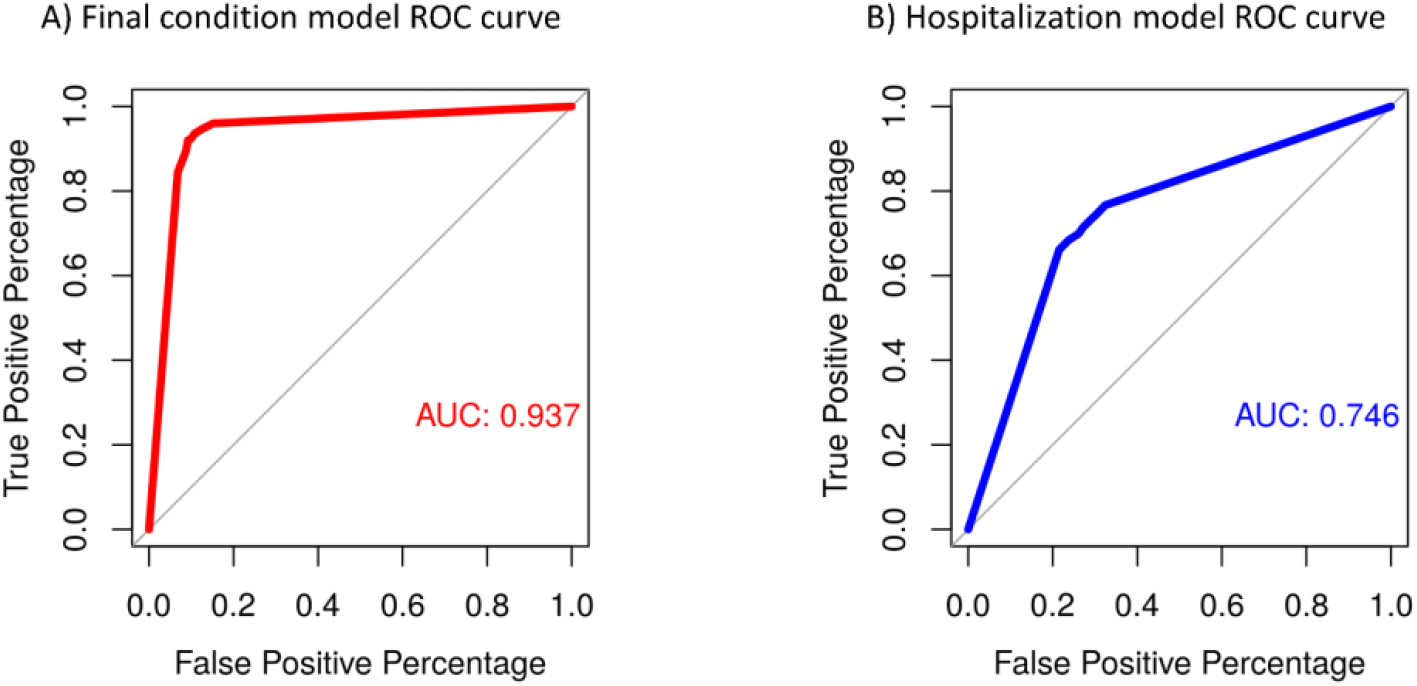
ROC curves for final condition and hospitalization predictive models.

**Supplementary Figure S4.**
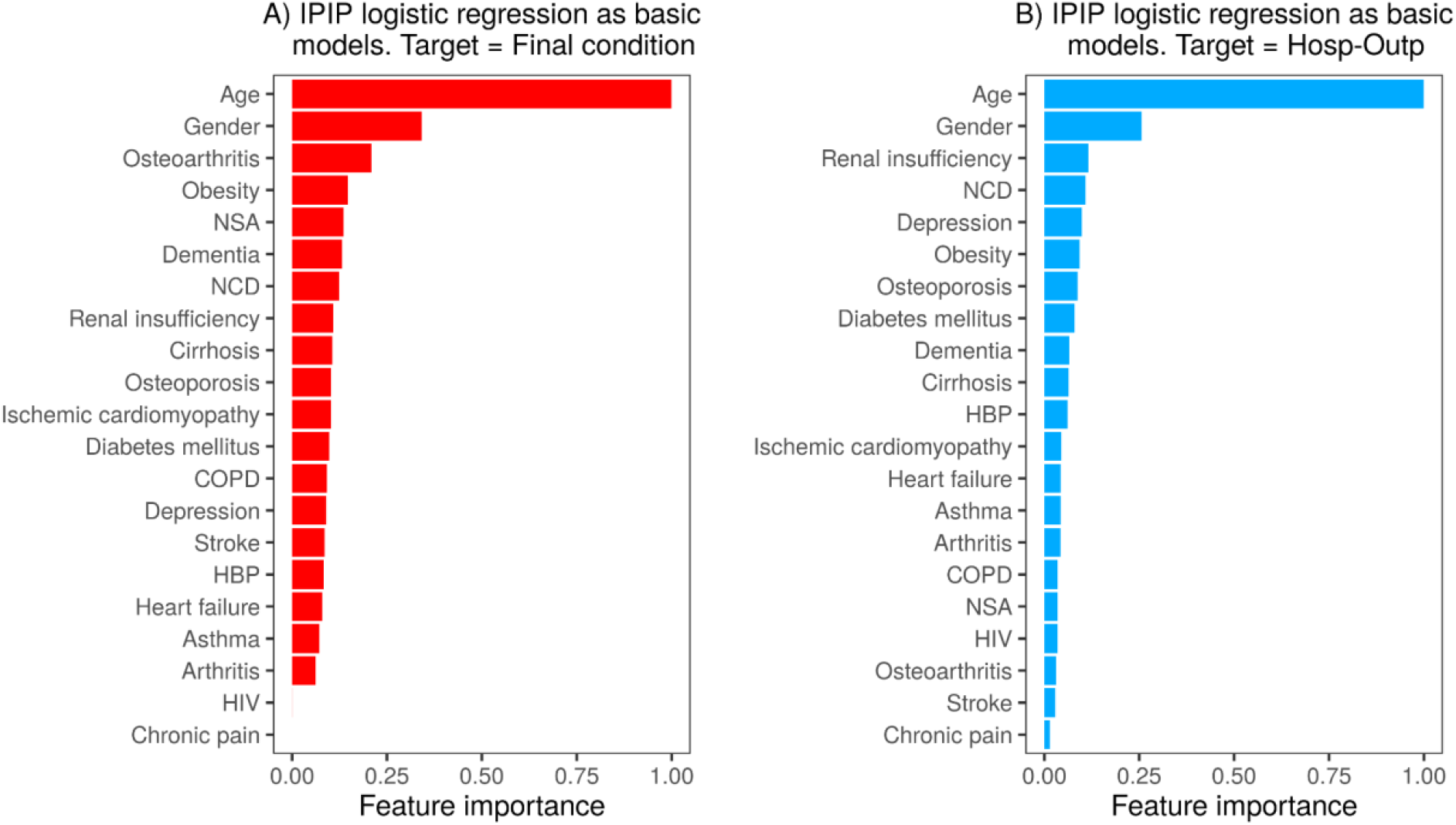
Features importance.

